# Allosensitization Status Predicts Excess Mortality in Congenital Heart Disease Transplant Recipients in the Current Era: An Analysis of the United Network for Organ Sharing Database

**DOI:** 10.1101/2025.02.07.25321899

**Authors:** Matthew J. O’Connor, Courtney Vu, Xuemei Zhang, Laura Bennett, Humera Ahmed, Jonathan J. Edwards, Kimberly Y. Lin, Yang Li, Katsuhide Maeda, Brooke Marcellus, Dimitrios Monos, Joseph W. Rossano, Carol A. Wittlieb-Weber, Jonathan B. Edelson

**Affiliations:** Division of Cardiology, Department of Pediatrics, Children’s Hospital of Philadelphia, University of Pennsylvania Perelman School of Medicine, Philadelphia PA USA; Data Science and Biostatistics Unit, Department of Biomedical and Health Informatics, Children’s Hospital of Philadelphia, Philadelphia PA USA; Immunogenetics Laboratory, Department of Pathology and Laboratory Medicine, Children’s Hospital of Philadelphia, University of Pennsylvania Perelman School of Medicine, Philadelphia PA USA; Division of Pediatric Cardiothoracic Surgery, Department of Surgery, Children’s Hospital of Philadelphia, University of Pennsylvania Perelman School of Medicine, Philadelphia PA USA

**Author notes:** Address for correspondence: Matthew J. O’Connor, MD Division of Cardiology Children’s Hospital of Philadelphia 3401 Civic Center Blvd Philadelphia, PA 19104.

## Abstract

**Background:** Allosensitization in pediatric heart transplantation (HT) is a challenging problem, with ongoing uncertainty as to optimal management strategy. Patients with congenital heart disease (CHD) have the highest risk of allosensitization and may be at risk for inferior outcomes following HT due to an accumulation of risk factors.

**Methods:** The United Network for Organ Sharing database was studied for all patients <18 years of age with CHD undergoing HT between April 2015 and December 2020. Patients were grouped into three categories of allosensitization status based on calculated panel reactive antibody (cPRA) obtained closest to the time of HT: nonsensitized (cPRA <10%), moderately sensitized (cPRA 10% - <80%), and highly sensitized (cPRA ≥80%). The primary outcome measures of interest were one-year patient and graft survival following HT. Multivariable analysis was used to control for differences in preoperative clinical characteristics among sensitization categories.

**Results:** During the study period, 1086 patients with CHD underwent HT at a median of 3 years of age. Nonsensitized patients comprised 70% of the cohort; 22% were moderately sensitized and 9% were highly sensitized. Unadjusted 1-year mortality was 25% in the highly sensitized group compared to 8.7% in the nonsensitized group (P<0.001). After adjustment, highly sensitized patients were >3 times more likely to die within the first year than nonsensitized patients (HR 3.44, 95% CI 2.13 - 5.54, P<0.001). The relationship between cPRA and crossmatch result was also assessed using multivariable regression. A variety of crossmatches were performed, including cytotoxicity and flow cytometry modalities. Regardless of crossmatch result, highly sensitized patients had an increased risk of one-year mortality and graft failure compared to nonsensitized and moderately sensitized patients (HR 3.4, 95% CI 1.98 – 5.84, P<0.001 and HR 3.32, 95% CI 1.94 – 5.67, P<0.001 for one-year mortality and the composite of death or graft failure, respectively).

**Conclusions:** Highly sensitized patients with CHD undergoing HT in the current era experience 25% 1-year mortality, which is significantly worse than less sensitized or nonsensitized patients. The magnitude of sensitization as reflected by cPRA, is highly predictive of adverse outcomes. These at-risk patients remain in need of more effective therapies for desensitization and management of the consequences of anti-HLA antibodies following HT.

**Clinical Perspective:** *What is New?:*

- Allosensitization to HLA antigens is a common problem in pediatric heart transplantation, and outcomes remain suboptimal in allosensitized patients undergoing heart transplantation. Patients with CHD are at the highest risk of allosensitization.
- In the current study, highly sensitized children with CHD undergoing heart transplantation in the current era experience 25% 1-year mortality following heart transplantation, which is significantly higher than in other groups undergoing transplantation.
- Allosensitization status, regardless of crossmatch result, independently predicted mortality following heart transplantation in this cohort.

*What are the Clinical Implications?:*

- Highly sensitized patients with CHD are much more likely to die in the first year following heart transplantation than less- or nonsensitized patients. They also experience higher rates of rejection, which contributes to morbidity and late mortality.
- Many efforts are made to minimize the likelihood of a positive crossmatch at the time of transplantation in order to optimize outcomes. However, the results of this study indicate that allosensitization status is the primary driver of outcomes when both allosensitization status and crossmatch result are taken into account. Therefore, continued development of new therapies for desensitization is warranted.

## Introduction

Allosensitization, also known as sensitization, is the state of having antibodies against non-self HLA antigens. In the context of pediatric HT, sensitization is most frequently observed as a consequence of exposure to allograft tissue utilized for surgical reconstruction in congenital heart disease (CHD) (1–3) but can also occur secondary to blood product exposure and VAD placement (3–6). Independent of mechanism, allosensitization is a risk factor for multiple adverse outcomes in pediatric HT, including prolonged waiting list times and mortality (7,8), increased risk of rejection (9), and increased risk of graft loss and mortality (8). While a number of therapies have been utilized to reduce the degree of allosensitization prior to HT (10,11), they are of variable efficacy, are accompanied by the risk of significant adverse effects, and may not work at all for some patients.

At the time of HT, the functional test for histocompatibility between the donor and recipient is embodied in the retrospective crossmatch, wherein recipient serum is tested against donor tissue to determine whether pre-formed anti-HLA antibodies have the capability of causing cell lysis in the graft (cytoxicity crossmatch), or merely bind to the graft (flow cytometry) (12). In general, avoidance of a positive cytotoxic and/or flow cytometry crossmatch is desired. Given the potential adverse outcomes associated with a positive crossmatch, the practical consequence of such an avoidance strategy is that many pediatric and adult HT programs will not offer HT to sensitized recipients when it is anticipated that the presence of sensitization will result in a positive crossmatch against a potential donor, although considerable variability exists among programs with respect to thresholds for what constitutes an unacceptable degree of sensitization and crossmatch result.

Because HT remains the only effective long-term therapy for end-stage heart failure (HF), some institutions will perform HT in highly sensitized candidates notwithstanding the aforementioned risks and have reported acceptable early outcomes (13, 14). In a recent prospective, multicenter study, HT in sensitized pediatric recipients with a positive cytotoxicity crossmatch has also been shown to be feasible with outcomes comparable to nonsensitized recipients (15), although the number of patients with a positive cytotoxicity crossmatch in this cohort was relatively small. Longer-term outcomes, morbidities such as rejection, and outcomes in unselected populations are less well known. Furthermore, the relationship between sensitization status and crossmatch result is complex: while nearly all patients with a positive crossmatch are sensitized, not all patients who are sensitized have a positive crossmatch, and it is unclear which factor is the primary driver of outcomes following HT in sensitized patients. As such, there continues to be uncertainty about the ideal approach to the highly sensitized pediatric HT recipient in the current era. We therefore sought to review the clinical characteristics and outcomes of sensitized pediatric HT recipients in the current era using the United Network for Organ Sharing (UNOS) database, focusing on outcomes based on the degree of sensitization and crossmatch status.

## Methods

Using the publicly available UNOS STAR dataset, we identified pediatric (<18 years of age) patients undergoing HT with a diagnosis of CHD in either the waitlist primary diagnosis or recipient primary diagnosis fields between 4/1/2015 and 12/31/2020. The diagnosis of CHD is listed in the UNOS database as “CHD with surgery” or “CHD without surgery,” without further specification as to the lesion. Patients undergoing multiple organ transplantation were excluded from the analysis. In addition, patients with missing data regarding the degree of sensitization (if any), crossmatch performed at the time of HT, covariates, or outcomes of interest were excluded from analysis. Follow-up was ascertained on all patients through 12/31/2021 to ensure all patients had at least one year of post-HT follow-up. The start date of 4/1/2015 was chosen because the reporting of allosensitization status in the UNOS database changed on that date and remained consistent moving forward; selection of this date also allowed for the creation of a contemporary cohort representing current practices in immunosuppression and immunomodulation. We focused specifically on patients with CHD given that an underlying diagnosis of CHD is the primary risk factor for allosensitization.

Allosensitization status was obtained by determining the calculated panel reactive antibody (cPRA) obtained closest to the time of HT. The cPRA is reported as a percentage, with a possible range of 0% to 100%, and encompasses class I and class II anti-HLA antibodies. Based on the cPRA value obtained closest to the time of HT, patients were grouped into three categories representing allosensitization status: nonsensitized (cPRA <10%); moderately sensitized (cPRA 10% - <80%), and highly sensitized (cPRA ≥80%). These cPRA cutoffs were created *a priori* and based on the generally accepted definition of sensitization as a cPRA ≥10% (16), as well as consensus opinion that cPRA ≥80% is a threshold above which desensitization therapy is usually considered (12). Crossmatch results were grouped according to the modality performed: flow cytometry and cytotoxic. Within each modality, results were also reported for T-cell and B-cell crossmatches. For all analyses, a positive crossmatch was defined as a positive crossmatch by flow cytometry and/or cytotoxicity. The primary outcome measures of interest were patient and graft survival at one year following HT. Secondary outcomes included the incidence of treated rejection prior to hospital discharge and treated rejection within one year following HT.

Summary data are presented as median with IQR or count with percentage as applicable. The Cochran-Armitage trend test was employed to assess the relationship between allosensitization status at the time of HT and crossmatch results.

For time-to-event outcomes (1-year mortality and 1-year graft failure or mortality) and rejection outcomes (acute rejection prior to hospital discharge and 1-year treated rejection post-HT), unadjusted analyses assessing the association of allosensitization status were first performed using the log-rank test or Cochran-Armitage trend test. A Kaplan-Meier plot was created to depict 1-year mortality across the three allosensitization groups. Next, adjusted analyses were performed using multivariable Cox regression or multivariable logistic regression. Covariates included in the models included age, mechanical ventilation at HT, ECMO at HT, graft ischemic time, dialysis prior to transplant, VAD at HT, and the year of HT. Backward selection and clinical judgment were used to build the multivariable models. The same multivariable regression models were used to evaluate the relationship between crossmatch results and outcomes. Finally, multivariable Cox regression models were employed to assess the individual associations between positive crossmatch and allosensitization status with death, graft failure, and rejection. This was performed in order to account for the contribution of allosensitization status to outcomes distinct from crossmatch result. All analyses were conducted using SAS version 9.4 (SAS Institute, Cary, NC). Kaplan-Meier plots were generated using the ggsurvfit package in RStudio version 2023.09.1 Build 494 (Posit Software, PBC, Boston, MA).

The UNOS STAR database is deidentified; therefore, local IRB approval was not required as this study did not meet the definition of human subjects research. The data reported here have been supplied by UNOS as the contractor for the Organ Procurement and Transplantation Network (OPTN). The interpretation and reporting of these data are the responsibility of the authors and in no way should be seen as an official policy of or interpretation by the OPTN or the U.S. Government. The data that support the findings of this study are available from the corresponding author upon reasonable request and after execution of a data use agreement. The first and senior authors of the manuscript (MJO, JBE) had full access to all of the data and the study and take responsibility for its integrity and the data analysis.

## Results

During the study period, 1352 pediatric patients with CHD and complete cPRA data underwent HT. cPRA data were missing in 266 patients; these patients were excluded so the analytic cohort therefore comprised 1086 patients. The demographic and clinical characteristics of these patients are shown in **Table 1**. At the time of HT, median age was 3 years (IQR 0 years – 11 years); 433 patients (39.9%) were female. Thirty-five patients (3.2%) were on ECMO at the time of HT and 160 patients (14.7%) were on a VAD. Mechanical ventilation and dialysis were utilized at the time of HT in 173 (15.9%) and 20 (1.8%) patients, respectively. Allosensitization status for the 1086 patients studied is also shown in **Table 1**. The majority of patients (N = 756, 70%) were nonsensitized; 22% (N = 234) were moderately sensitized (cPRA 10 – 79%); 9% (N = 96) were highly sensitized (cPRA ≥80%).

**Table 1.** Demographic and Clinical Characteristics of the Population (N = 1086)

| Characteristic | Median (IQR) or N (%) |
| --- | --- |
| Age at HT | 3 (0 -11) years |
| Female sex | 433 (39.9) |
| Mechanical ventilation at HT | 173 (15.9) |
| ECMO at time of HT | 35 (3.2) |
| VAD at time of HT | 160 (14.8) |
| Dialysis at time of HT | 20 (1.8) |
| Allosensitization status at time of HT |  |
| Nonsensitized (cPRA <10%) | 756 (69.6) |
| Moderately sensitized (cPRA 10-79%) | 234 (21.5) |
| Highly sensitized (cPRA ≥80%) | 96 (8.8) |

The interaction between allosensitization status at the time of HT and crossmatch result is shown in **Table 2**. The majority of patients (86%) had a flow cytometry crossmatch (T and/or B cell) reported. Cytotoxicity crossmatches (T and/or B cell) were less commonly performed overall (27% of cohort) but increased in frequency for patients as the magnitude of sensitization increased. For example, 18% of nonsensitized patients had a B cell cytotoxicity crossmatch performed, while 34% of highly sensitized patients had a B cell cytotoxicity crossmatch performed (P = 0.0005). As expected, the incidence of positive crossmatch (by flow cytometry or cytotoxicity) increased as the magnitude of sensitization increased. For example, in nonsensitized patients (cPRA <10%) the incidence of positive T cell flow cytometry and T cell cytotoxicity crossmatch was 3% and 4%, respectively. For moderately sensitized patients, a positive T cell flow cytometry crossmatch was seen in 16% and a positive T cell cytotoxicity crossmatch was seen in 10%. In highly sensitized patients, a positive T cell flow cytometry crossmatch was seen in 46% of patients and a positive T cell cytotoxicity crossmatch was seen in 24% of patients (P < 0.0001 for trend). A minority (16.4% of the entire cohort) had both flow cytometry and cytotoxicity crossmatches performed, although this proportion increased to 27.1% for highly sensitized patients.

**Table 2.** Relationship Between cPRA and Crossmatch Result at HT (N = 1086)

|  | Total | Nonsensitized<br>(cPRA<10%) | Moderately<br>sensitized<br>(cPRA<br>10%-<80%) | Highly<br>sensitized<br>(cPRA<br>≥80%) | P value |
| --- | --- | --- | --- | --- | --- |
| N | 1086 | 756 | 234 | 96 | - |
| B cell flow cytometry |  |  |  |  |  |
| Flow cytometry performed | 928 (85.4) | 637 (84.3) | 209 (89.3) | 82 (85.4) | 0.16 |
| Flow cytometry positive | 135 (14.5) | 38 (5.9) | 50 (21.4) | 47 (57.3) | <0.0001 |
| T cell flow cytometry |  |  |  |  |  |
| Flow cytometry performed | 934 (86.0) | 640 (84.7) | 210 (89.7) | 84 (87.5) | 0.13 |
| Flow cytometry positive | 95 (10.2) | 22 (3.4) | 34 (16.2) | 39 (46.4) | <0.0001 |
| B cell cytotoxicity |  |  |  |  |  |
| Cytotoxicity performed | 223 (20.5) | 136 (18.0) | 54 (23.1) | 33 (34.4) | 0.0005 |
| Cytotoxicity positive | 33 (14.8) | 9 (6.6) | 8 (14.8) | 16 (48.5) | <0.0001 |
| T cell cytotoxicity |  |  |  |  |  |
| Cytotoxicity performed | 291 (26.8) | 188 (24.9) | 70 (29.9) | 33 (34.4) | 0.07 |
| Cytotoxicity positive | 22 (7.6) | 7 (3.7) | 7 (10.0) | 8 (24.2) | 0.0001 |
| At least one crossmatch modality<br>performed | 1060 (97.6) | 733 (97.0) | 231 (98.7) | 96 (100) | 0.0266 |
| Both flow cytometry and<br>cytotoxicity crossmatch<br>performed | 178 (16.4) | 98 (13.0) | 54 (23.1) | 26 (27.1) | <0.001 |

Data are presented as N (%)

Kaplan-Meier analysis was performed to assess the relationship between allosensitization status at the time of HT and the outcome of death or graft failure within the first year after HT. The results of this analysis are shown in **Figure 1**. In nonsensitized patients, 1-year mortality was seen in 8.7%, while in highly sensitized patients, 25% mortality was encountered in the first year following HT (P<0.001) (**Table 3**). For rejection outcomes, 10.9% of nonsensitized patients were treated for acute rejection prior to hospital discharge, increasing to 17.5% for moderately sensitized patients and 33.3% for highly sensitized patients (P<0.001). In the first year post-HT, 16.8% of nonsensitized patients, 24.1% of moderately sensitized patients, and 33.8% of highly sensitized patients were treated for rejection, respectively (P<0.001).

**Figure 1.**
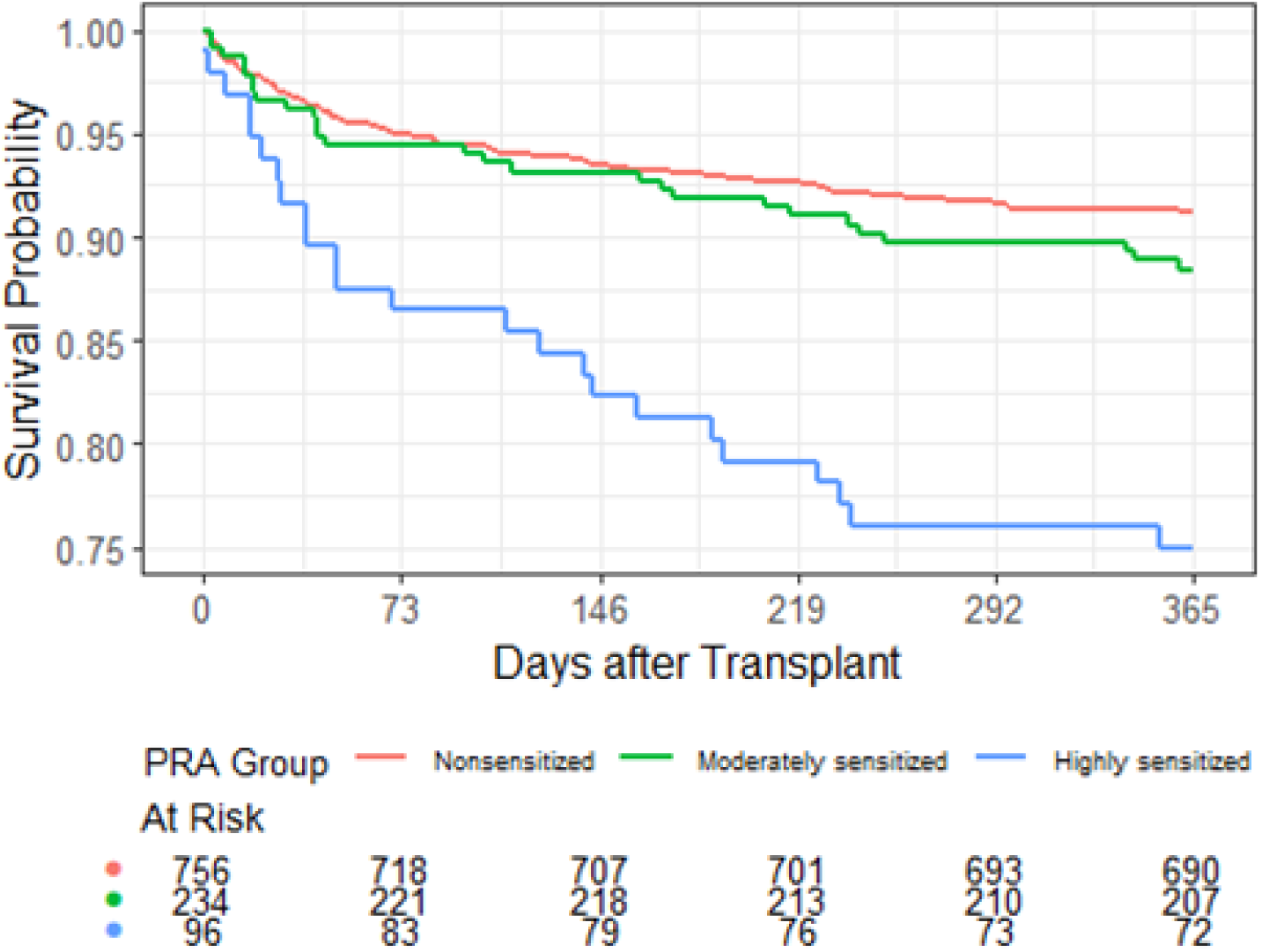
Kaplan-Meier plot evaluating 1-year mortality in patients stratified by allosensitization status.

**Table 3.** Relationship Between Allosensitization Status at Time of HT and Outcomes of 1-Year Mortality, Graft Failure, and Rejection

| Percentage of Failure<br>From Kaplan Meier Estimate | Nonsensitized<br>(cPRA<10%) | Moderately<br>sensitized<br>(cPRA<br>10%-<80%) | Highly<br>sensitized<br>(cPRA ≥80%) | P<br>value* |
| --- | --- | --- | --- | --- |
| 1 year mortality after HT | 8.7% | 11.5% | 25.0% | <0.001 |
| Graft failure or mortality in first year<br>post-HT<sup>†</sup> | 9.0% | 11.6% | 25.0% | <0.001 |
| Treated for acute rejection prior to<br>hospital discharge | 10.9% | 17.5% | 33.3% | <0.001 |
| Treated for rejection in first year post-HT** | 16.8% | 24.1% | 33.8% | <0.001 |

\*p-value of Log rank test for mortality and graft failure in first year after transplant, and
Cochran-Armitage Trend test for rejection outcomes
<sup>†</sup>The number of patients with missing graft failure information is 3.
\*\* The number of patients with missing one year rejection information is 162.

The relationship between allosensitization status at the time of HT and the likelihood of death or graft failure was also assessed via multivariable Cox regression, as shown in **Table 4**. Highly sensitized patients were greater than 3 times more likely to die within the first year post-HT than nonsensitized patients (HR 3.44, 95% CI 2.13 - 5.54, P<0.001) after adjustment for baseline clinical characteristics; there was no survival difference between nonsensitized and moderately sensitized groups. For the secondary outcomes of acute rejection prior to hospital discharge or within the first year post-HT, moderately sensitized and highly sensitized patients had a higher likelihood of both secondary outcomes, with highly sensitized patients seeing the greatest risk (**Table 5**). The relationship between crossmatch result and the likelihood of death or graft failure was also examined using multivariable Cox regression, shown in **Table 6**. Patients with a positive crossmatch (of any type) had an increased likelihood of mortality in the first year after HT compared to those with a negative crossmatch (HR 1.63, 95% CI 1.06 – 2.52, P = 0.027) **(Figure 2)**, and also had an increased risk of rejection prior to discharge as well as within the first year post-HT (OR 2.55, 95% CI 1.71 – 3.81, P<0.001) (**Table 7**).

**Table 4.** Relationship Between Allosensitization Status and 1-year post-HT Mortality and Graft Failure Using Multivariable Cox Regression

|  | Hazard Ratio | 95% CI | P value |
| --- | --- | --- | --- |
| **1 year mortality after HT\*** |  |  |  |
| Nonsensitized (cPRA <10% (ref.)) | - | - | - |
| Moderately sensitized (cPRA 10%-<80%) | 1.41 | 0.90-2.21 | 0.13 |
| Highly sensitized (cPRA ≥80%) | 3.44 | 2.13-5.54 | <0.001 |
| **Graft failure or mortality in 1 year after HT <sup>†</sup>** |  |  |  |
| Nonsensitized (cPRA <10% (ref.)) | - | - | - |
| Moderately sensitized (cPRA 10%-<80%) | 1.37 | 0.87-2.14 | 0.17 |
| Highly sensitized (cPRA ≥80%) | 3.34 | 2.08-5.37 | <0.001 |

\* The number of patients with missing covariates is 1.
<sup>†</sup>The number of patients with missing covariates is 3.
The variables adjusted in the death models and graft failure model are age, mechanical ventilation at transplant, ECMO at transplant, ischemic time, dialysis prior to transplant, VAD at transplant, and year of transplant.

**Table 5:** Relationship Between Allosensitization Status and Secondary Outcomes (Rejection Prior to Discharge or Within First Year Post-HT) Using Multivariable Logistic Regression

|  | Odds Ratio | 95% CI | P value |
| --- | --- | --- | --- |
| **Treated for acute rejection prior to hospital discharge\*** |  |  |  |
| Nonsensitized (cPRA <10% (ref.)) | - | - | - |
| Moderately sensitized (cPRA 10%-<80%) | 1.78 | 1.18 - 2.69 | 0.006 |
| Highly sensitized (PRA ≥80%) | 4.28 | 2.61 - 7.03 | <0.001 |
| **Treated for rejection in first year post-transplant<sup>†</sup>** |  |  |  |
| Nonsensitized (cPRA <10% (ref.)) | - | - | - |
| Moderately sensitized (cPRA 10%-<80%) | 1.58 | 1.07 - 2.32 | 0.020 |
| Highly sensitized (cPRA ≥80%) | 2.57 | 1.47 - 4.48 | <0.001 |

\*The number of patients with missing covariates is 1.
<sup>†</sup>The number of patients with missing one year rejection information is 163.
The variables adjusted in the model are age, mechanical ventilation at transplant, ECMO at transplant, ischemic time, dialysis prior to transplant, VAD at transplant, and year of transplant.

**Table 6:** Relationship between Positive Crossmatch and Mortality/Graft Failure One Year Following HT Using Multivariable Cox Regression (N = 1060; patients with no crossmatch reported excluded from analysis)

|  | Hazard Ratio | 95% CI | P value |
| --- | --- | --- | --- |
| 1 year mortality after HT* |  |  |  |
| Negative crossmatch | 1.00 | - | - |
| Positive crossmatch | 1.63 | 1.06 - 2.52 | 0.027 |
| Death or graft failure in first-year after HT<sup>†</sup> |  |  |  |
| Negative crossmatch | 1.00 | - | - |
| Positive crossmatch | 1.59 | 1.03 - 2.46 | 0.036 |

\*The number of patients with missing covariates excluded from the analysis is 1.
<sup>†</sup>The number of patients with missing covariates excluded from the analysis is 4.
The variables adjusted in the model are age, mechanical ventilation at transplant, ECMO at transplant, ischemic time, dialysis prior to transplant, VAD at transplant, and year of transplant.

**Figure 2.**
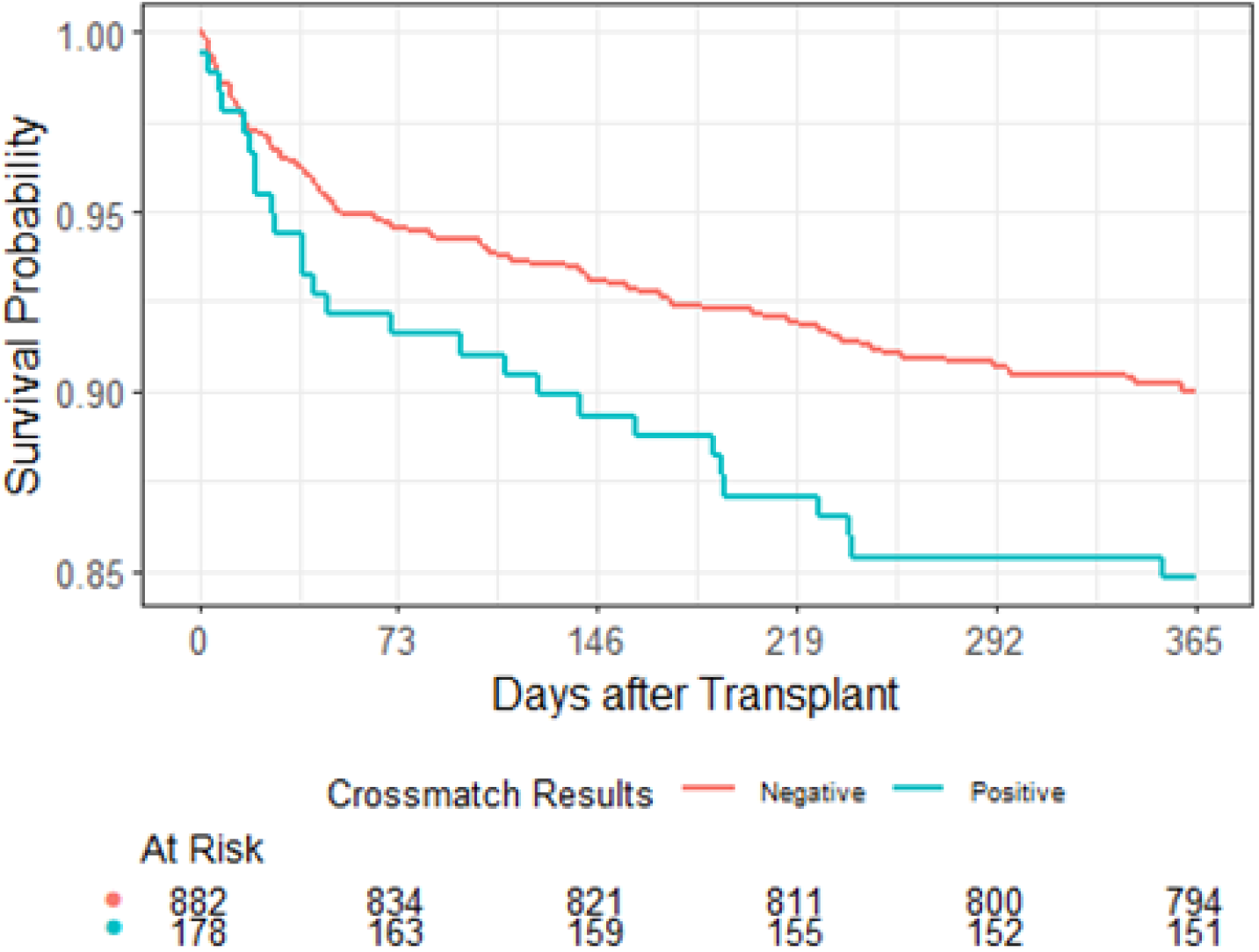
Kaplan-Meier plot evaluating 1-year mortality in patients stratified by crossmatch result.

**Table 7:** Relationship between Positive Crossmatch and Acute Rejection Prior to Discharge or Within First Year After HT using Multivariable Logistic Regression (N = 1060, patients with no crossmatch reported excluded from analysis)

|  | Odds Ratio | 95% CI | P value |
| --- | --- | --- | --- |
| Treated for acute rejection prior to hospital discharge* |  |  |  |
| Negative crossmatch | 1.00 | - | - |
| Positive crossmatch | 2.55 | 1.71 - 3.81 | <.001 |
| Treated for rejection in first year after HT<sup>†</sup> |  |  |  |
| Negative crossmatch | 1.00 | - | - |
| Positive crossmatch | 1.69 | 1.11 - 2.57 | 0.014 |

\* The number of patients with missing covariates excluded from the analysis is 1.
<sup>†</sup> The number of patients with missing covariates excluded from the analysis is 161.
The variables adjusted for are: age, mechanical ventilation at transplant, ECMO at transplant, ischemic time, dialysis prior to transplant, VAD at transplant, and year of transplant.

Finally, the relationships between sensitization status and crossmatch result with one-year mortality, graft failure, and rejection were evaluated using multivariable Cox. After adjusting for allosensitization status, there was no difference in one-year mortality after HT between negative and positive crossmatch patients; however, highly sensitized patients had an increased risk of one-year mortality and graft failure compared to nonsensitized and moderately sensitized patients (HR 3.4, 95% CI 1.98 – 5.84, P<0.001 and HR 3.32, 95% CI 1.94 – 5.67, P<0.001 for one-year mortality and death or graft failure, respectively), irrespective of the crossmatch result. Each possible combination of crossmatch result (positive or negative) and pre-HT cPRA category (nonsensitized, moderately sensitized, highly sensitized) was evaluated. This demonstrated that only highly sensitized patients, regardless of crossmatch result, had worse post-HT outcomes. These findings are summarized in **Table 8**. Comparable outcomes for the secondary outcomes of treated rejection either prior to hospital discharge or in the first year post-HT were seen and are presented in **Supplemental Table 1**. One important difference in the secondary outcomes, in contrast to the primary outcome, is that a positive crossmatch was associated with an increased risk of treated rejection prior to hospital discharge, even after accounting for allosensitization status.

**Table 8:**
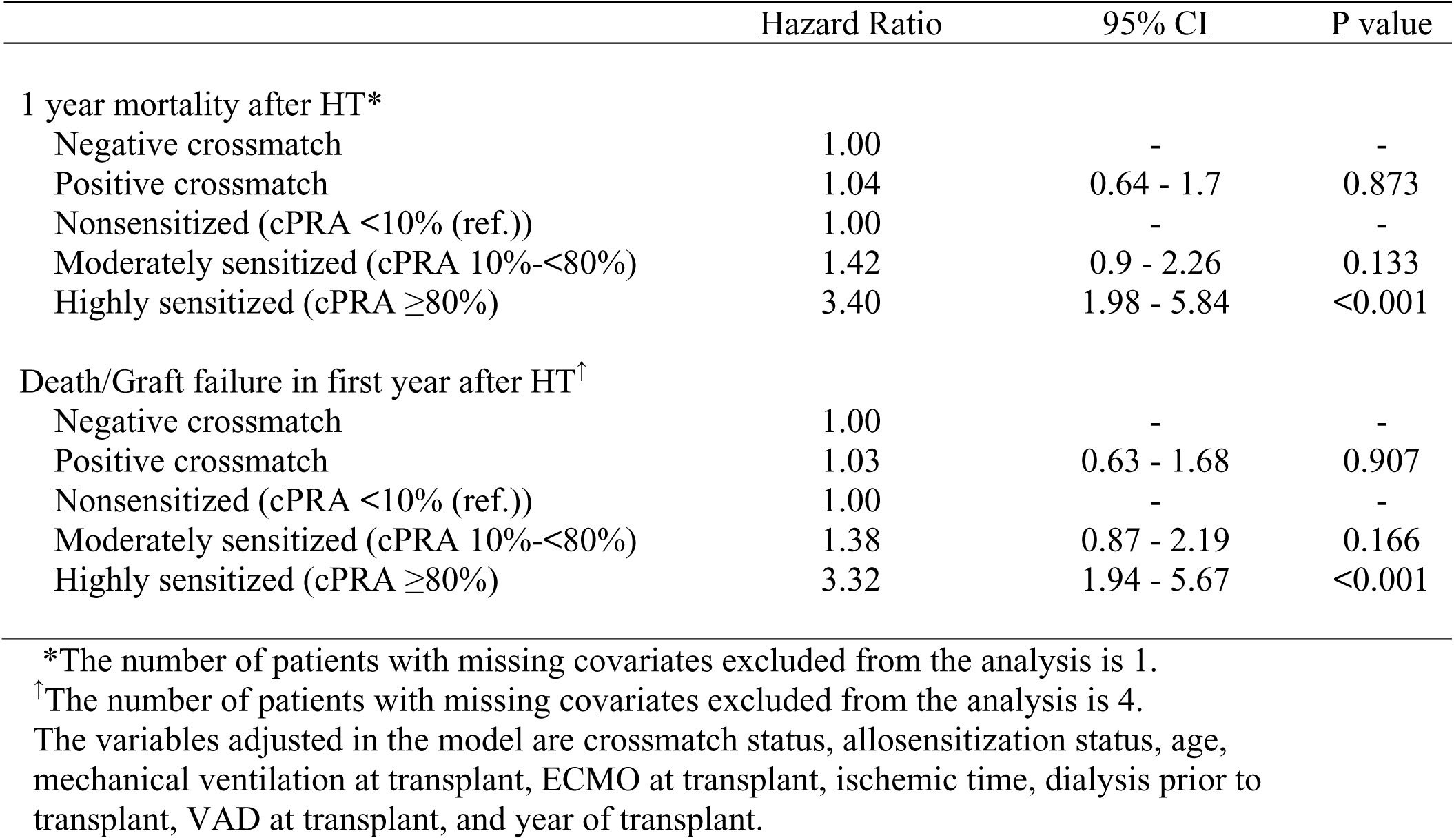
Multivariable Cox Regressions to Assess the Relationship between Positive Crossmatch and Allosensitization Status with Death/Graft Failure (N = 1060; patients with no crossmatch reported excluded from analysis)

|  | Hazard Ratio | 95% CI | P value |
| --- | --- | --- | --- |
| 1 year mortality after HT* |  |  |  |
| Negative crossmatch | 1.00 | - | - |
| Positive crossmatch | 1.04 | 0.64 - 1.7 | 0.873 |
| Nonsensitized (cPRA <10% (ref.)) | 1.00 | - | - |
| Moderately sensitized (cPRA 10%-<80%) | 1.42 | 0.9 - 2.26 | 0.133 |
| Highly sensitized (cPRA ≥80%) | 3.40 | 1.98 - 5.84 | <0.001 |
| Death/Graft failure in first year after HT<sup>†</sup> |  |  |  |
| Negative crossmatch | 1.00 | - | - |
| Positive crossmatch | 1.03 | 0.63 - 1.68 | 0.907 |
| Nonsensitized (cPRA <10% (ref.)) | 1.00 | - | - |
| Moderately sensitized (cPRA 10%-<80%) | 1.38 | 0.87 - 2.19 | 0.166 |
| Highly sensitized (cPRA ≥80%) | 3.32 | 1.94 - 5.67 | <0.001 |

\*The number of patients with missing covariates excluded from the analysis is 1.
<sup>†</sup>The number of patients with missing covariates excluded from the analysis is 4.
The variables adjusted in the model are crossmatch status, allosensitization status, age, mechanical ventilation at transplant, ECMO at transplant, ischemic time, dialysis prior to transplant, VAD at transplant, and year of transplant.

## Discussion

Management of the sensitized patient being considered for HT continues to be a formidable challenge, with a robust literature illustrating the sustained efforts within the field to optimally treat this complex patient population. In the present analysis of the UNOS dataset examining a contemporary cohort of pediatric patients with CHD undergoing HT, we examined outcomes according to sensitization and crossmatch status, with a focus on attempting to clarify the relationship between sensitization status and crossmatch result. Several important findings emerge from our analysis. First, nearly 30% of patients in this cohort underwent HT with a cPRA of 10% or higher; a smaller proportion (9%) were highly sensitized with a cPRA of ≥80%. Second, the group of highly sensitized patients (cPRA ≥80%) had very high mortality in the first year post-HT (25%), with at least one-third of this group being treated for rejection in the first year-post transplant. Third, after controlling for baseline clinical characteristics as well as allosensitization status, a positive crossmatch did not independently predict mortality or graft loss in the first year following HT. Only the highly sensitized group experienced significantly increased mortality after HT, regardless of crossmatch result. Taken together, this suggests that pre-HT sensitization status is a stronger predictor of post-HT outcome than crossmatch result in patients with CHD.

Highly sensitized patients have consistently been shown to have suboptimal outcomes following HT, even in relatively recent cohorts of HT recipients. These patients are at higher risk of hyperacute rejection, acute rejection, antibody-mediated rejection, coronary allograft vasculopathy, and ultimately demonstrate poorer survival than nonsensitized recipients (8, 17–18). Various strategies to reduce anti-HLA antibodies have been employed, including pre-HT “desensitization” therapies, peri-HT antibody depletion through plasmapheresis, and intensive immunosuppression strategies following HT, including prolonged corticosteroid administration. Such therapies are of variable benefit and may have significant side effects, not to mention increased cost and overall healthcare utilization.

Our work builds upon prior efforts in the field to understand the implications of sensitization on outcomes following HT. One of the many difficulties involved in managing a highly sensitized patient at the time of HT is integrating the results of the pre-HT antibody testing (embodied in the cPRA) with the post-HT histocompatibility testing (embodied in the retrospective crossmatch). Changing methodologies of anti-HLA antibody assessment and crossmatch testing further complicate clinical decision-making. Although so-called “virtual crossmatching” permits clinicians to determine the likelihood of a positive crossmatch with a potential donor based on the HLA typing of the donor and the antibody specificities of the recipient, it is not a foolproof method and much of the decision-making when interpreting a virtual crossmatch is whether the results of such an analysis predict a positive flow or cytotoxic crossmatch (or both). Given these uncertainties, pre-HT antibody testing remains important in establishing a potential HT recipient’s degree of immunologic risk.

In a prior analysis of the UNOS database from our group, we evaluated changes in the methodology of pre-HT anti-HLA antibody assessment in a population of pediatric and adult HT recipients and found an increase in the proportion of sensitized patients in the relatively recent era, which was coincident with an increase in the utilization of flow cytometric techniques for anti-HLA antibody assessment (19). In this aforementioned study, we also found that a pre-HT cPRA that was positive (≥10%) by flow cytometry independently predicted graft loss after multivariable adjustment. In the present study, we also found that highly sensitized patients, regardless of crossmatch result, had worse outcomes, including rejection and mortality in addition to graft loss. When viewed from the perspective of the crossmatch, recent work from our group also demonstrated that patients with a positive flow cytometric crossmatch did not have worse graft survival than patients with a negative crossmatch (either by flow cytometry or cytotoxicity); only patients with a positive cytotoxic crossmatch had worse graft survival (20). These findings were confirmed in a recent study of pediatric HT recipients between 1999 – 2019 by Milligan et al, in which only patients with a crossmatch positive by cytotoxicity had a higher likelihood of graft loss, death, and treated rejection (21). Importantly, this latter study included patients with all diagnoses leading to HT, including CHD and cardiomyopathy. What distinguishes the results of the current study from these prior investigations is the evaluation of post-HT outcomes taking into account both allosensitization status and crossmatch result in a contemporary cohort. While pre-HT sensitization status and crossmatch result have both been individually associated with higher post-HT mortality risk, the results of the current study suggest that pre-HT sensitization status is a more powerful predictor of post-HT outcomes in children with CHD. This finding has obvious relevance to clinicians, since the sensitization status is known prior to HT and can therefore be used to predict post-HT outcome and consider alternatives to HT, if feasible, whereas the crossmatch result is generally only known after the HT has occurred, unless a prospective crossmatch is performed.

In the cohort of children with CHD undergoing HT in the present analysis, the 1-year mortality of 25% observed in patients with a cPRA ≥80% is a cause for concern. While several centers and multicenter collaborations have reported on acceptable early outcomes in highly sensitized patients with a positive cytotoxic crossmatch (13–15), these findings suggest that such outcomes may not be generalizable to the entire pediatric population undergoing HT and highlight that the subpopulation of sensitized patients with CHD are at particularly high risk of poor post-HT outcomes. Although beyond the scope of the present analysis, patients with CHD often come to transplant in a frail state (22) and are less likely to be supported with VADs at the time of listing for HT (23), the latter of which is an important tool in facilitating rehabilitation in patients of all ages with advanced heart failure.

It is likely that the high mortality observed in the highly sensitized group results from an accumulation of morbidities, of which antibody-mediated rejection (and the complications incurred from the therapies administered to treat it) play an important part. Although chronic therapy with dischargeable, durable VADs remains uncommon in children, the high post-HT mortality observed in the highly sensitized group should prompt consideration of employing such support when feasible and after a careful discussion of the risks and benefits with the patient’s family. Continued improvements in patient selection and surgical technique have resulted in improved survival following VAD implantation in children, even for those with complex CHD, including single ventricle circulations. Another alternative to high-risk HT in very highly sensitized patient is combined heart-liver transplantation; small series of combined heart-liver transplantation in highly sensitized patients have reported promising outcomes from the standpoint of survival, rejection, and donor-specific antibody production, likely owing to the transplanted liver’s ability to sequester and neutralize circulating anti-HLA antibodies (24). However, combined heart-liver transplantation is practically challenging and may raise concerns regarding the fairness of organ distribution when considering it in patients without intrinsic liver disease; the latter concern may be mitigated by considering “domino” transplantation of the recipient native liver (25).

Finally, while the findings from this study should prompt caution when considering HT in a highly sensitized patient with CHD, the fact remains that HT is the definitive therapy for end-stage heart failure and provides the best opportunity for long-term and sustained improvements in quality of life in this high-risk population. While 1-year post-HT mortality approximating 25% is well in excess of the mortality experienced in lower-risk groups, for many patients and families, this risk may be acceptable when compared against the near-certain risk of mortality for end-stage CHD. The findings from this study should continue to prompt careful examination of risk adjustment models used by regulatory agencies and other outcomes monitoring bodies with the goal of maximizing the benefit of HT to the candidates most in need of this therapy.

## Limitations and Future Directions

There are several important limitations to this study. While the strength of the UNOS dataset lies in the fact that it captures all of the HTs performed in the US with excellent follow-up data, some specificity is lacking with respect to underlying diagnoses leading to HT, practice variation regarding management of the sensitized HT candidate, and HLA antibody specificities that may influence outcomes in either a favorable or unfavorable direction. Missing data were uncommon for crossmatch results, but there was increased missingness with respect to rejection follow-up. In addition, patients older than 18 years of age were not evaluated in this study. There was also significant variation in the types of crossmatch performed at the time of HT. Flow cytometric crossmatches were much more commonly performed than cytotoxic crossmatches, even in highly sensitized patients. This may explain why sensitization predicted mortality whereas crossmatch status did not. Finally, cPRA is a broad measure of an individual’s immunologic risk prior to HT and does not take into account the influence of individual antibody specificities; for example, a patient with a low cPRA may nonetheless possess certain donor-specific antibodies at high titer that could prove problematic following HT. Our results should therefore not be construed to imply that all patients with cPRA <80% are immunologically low-risk, and, conversely, that all patients with a cPRA ≥80% are high risk.

In our center, based on our previous work (20), we perform a pre-HT virtual and/or prospective cytotoxicity crossmatch and proceed with HT if the cytotoxicity crossmatch is negative. For future study, we plan to examine those cases with cytotoxicity negative crossmatches and assess outcomes, including donor specific antibody (DSA) production; DSA information is not available in the UNOS dataset. Assessing the influence of DSA in negative cytotoxicity crossmatch cases will probably reveal a threshold level of HLA antibodies pre-HT (that become DSA post-HT) that would suggest a negative cytotoxicity crossmatch. Therefore, it could be used as a criterion for transplanting patients with cPRA ≥80% by supplementing the cPRA sensitization profile with anticipated DSA post-HT and provide an improved algorithm for increasing access to HT in highly sensitized patients.

## Conclusions

In a contemporary cohort of pediatric patients with CHD, highly sensitized recipients with a cPRA of ≥80% at the time of HT had a 3-fold increase in post-HT mortality or graft loss when compared to less sensitized recipients. Outcomes for these high-risk patients remain suboptimal and highlight the ongoing need for improvement in preventing and treating sensitization.

## Data Availability

The data that support the findings of this study are available from the corresponding author upon reasonable request and after execution of a data use agreement. The first and senior authors of the manuscript (MJO, JBE) had full access to all of the data and the study and take responsibility for its integrity and the data analysis.

## Acknowledgments

None

## Non-standard Abbreviations and Acronyms

HT: heart transplantation
CHD: congenital heart disease
cPRA: calculated panel reactive antibody
UNOS: United Network for Organ Sharing
OPTN: Organ Procurement and Transplantation Network
DSA: Donor-specific antibody

## Supplemental Publication Material

**Supplemental Table 1:** Multivariable Logistic Regressions to Assess Relationship Between Positive Crossmatch and Allosensitization Status with Acute Rejection Prior to Discharge/Treated for Rejection in First Year After HT (N = 1060; patients with no crossmatch reported excluded from analysis)

|  | Odds Ratio | 95% CI | P value |
| --- | --- | --- | --- |
| Treated for acute rejection prior to hospital discharge* |  |  |  |
| Negative crossmatch | 1.00 | - | - |
| Positive crossmatch | 1.64 | (1.04 - 2.59) | 0.035 |
| PRA <10% (ref.) | 1.00 | - | - |
| PRA 10%-<80% | 1.66 | (1.08 - 2.55) | 0.020 |
| PRA ≥80% | 3.32 | (1.9 - 5.81) | <.001 |
| Treated for rejection in first year after HT<sup>†</sup> |  |  |  |
| Negative crossmatch | 1.00 | - | - |
| Positive crossmatch | 1.3 | (0.82 - 2.05) | 0.263 |
| PRA <10% (ref.) | 1.00 | - | - |
| PRA 10%-<80% | 1.53 | (1.03 - 2.29) | 0.036 |
| PRA ≥80% | 2.27 | (1.25 - 4.14) | 0.007 |

\* The number of patients with missing covariates excluded from the analysis is 1.
<sup>†</sup> The number of patients with missing covariates excluded from the analysis is 161.
The variables adjusted for are crossmatch status, allosensitization status, age, mechanical ventilation at transplant, ECMO at transplant, ischemic time, dialysis prior to transplant, VAD at transplant, and year of transplant.

